# Elevated blood glucose levels are associated with the progression of brain hypometabolism, and HDL-C and *APOE4* add to this association

**DOI:** 10.1101/2024.09.20.24314082

**Authors:** Aravind Lathika Rajendrakumar, Svetlana Ukraintseva, Olivia Bagley, Matt Duan, Anatoliy I. Yashin, Konstantin G. Arbeev, the Alzheimer’s Disease Neuroimaging Initiative

## Abstract

**Background:** Brain glucose hypometabolism has consistently been found in neurodegenerative disorders, including Alzheimer’s disease (AD). High blood glucose and HDL cholesterol (HDL-C) levels have also been linked to neurodegeneration and AD. However, there is limited understanding of the relationships between dementia-related risk factors in the brain and blood.

**Methods:** A linear mixed model was used to examine the relationship between blood glucose and HDL-C levels and the progression of brain hypometabolism, adjusting for *APOE4* and other clinical covariates. The hypometabolic convergence index (HCI) was measured by fluorodeoxyglucose-18 (FDG) positron emission tomography (PET) in participants from the Alzheimer’s Disease Neuroimaging Initiative (ADNI). Data visualizations were generated to understand the joint effects of plasma glucose, HDL-C, and *APOE4* on HCI.

**Results:** There were 336 individuals (781 observations), of whom 22.62% had AD. The majority were male (63.98%) and of white race, and 48.51% were carriers of *APOE4*. Over time, high blood glucose level was associated with the progression of brain glucose hypometabolism (β=0.33, 95% CI: 0.02, 0.64, p<0.05). A high plasma HDL-C level (β=1.22, 95% CI: 0.09, 2.35, p<0.05), more study visits (β=1.67, 95% CI: 1.37, 1.98, p<0.001), and being an *APOE4* allele carrier (β=1.29, 95% CI: 0.15, 2.42, p<0.05) were also significant predictors of brain hypometabolism progression. *APOE4* carrier status and number of visits account for the largest proportion of the variance from the fixed effects model. Random effects due to participant characteristics and fixed effects together accounted for 95.2% of the model variance. Subgroup analysis revealed that these effects were observed only in those without AD.

**Conclusion:** High plasma glucose levels facilitated the progression of brain hypometabolism. The effect was more prominent in the *APOE4* double-carriers with elevated HDL-C. Elevated blood glucose may reflect systemic insulin resistance, which could impair brain glucose uptake, resulting in brain hypometabolism. Controlling blood glucose and HDL-C levels in *APOE4* carriers may improve brain metabolism, potentially delaying the onset of dementia.

## Background

Glucose usage in the brain has been finely tuned throughout evolution.^1^ Brain cells are highly energy-dependent and require a constant supply of glucose for optimal functioning.^2^ Glucose meets the energy demands for a diverse range of activities, including brain signaling, neurotransmitter production, and maintaining homeostasis.^3^ Due to this, multilayered mechanisms, including sensors, glucose transporters, enzymes, and specific cell pathways, work together to ensure the availability of glucose.^2^ Complex learning processes in neurons and astrocytes are correlated with brain metabolism, which is directly dependent on glucose usage.^4^ Exposure to insufficient glucose supply can harm memory and learning, and prolonged insufficiency can potentially cause permanent brain alterations.^3^ Thus, any deviation from the normal glucose uptake pattern in the brain might indicate a serious medical illness.^5^

Earlier neuroimaging studies have considerably expanded our knowledge of metabolic alterations in dementia.^6,7^ Neuroimaging outcomes are superior to traditional cognitive assessments in detecting related changes and correlate well with neuropathological changes in individuals with dementia.^8^ A pattern of glucose deficit in the brain is noted for several neurodegenerative disorders, which can be distinguished among the relevant conditions.^9^ In the case of Alzheimer’s disease (AD), hypometabolism begins much earlier than the actual onset of clinical symptoms and contributes to the further progression of the disease.^10^ The [^18^F]-2-fluoro-2-deoxy-2-glucose (FDG) tracer-based positron emission tomography (PET) is a widely used diagnostic method to ascertain the metabolic rate in different tissues. FDG mimics glucose absorption and remains in the body longer than glucose. Studying its buildup in tissues helps quantify the metabolic rate.^11^ The FDG PET-derived hypometabolic convergence index can accurately distinguish the AD signature brain hypometabolic pattern through automated brain image analysis.

Dementia subtypes, such as AD, have a complex, multifactorial etiology, stemming from an interplay between aging, genetics, and the environment.^12^ In this context, among the risk factors, hyperglycemia, and particularly diabetes, are major concerns due to its rising global prevalence.^13^ Clinical features of diabetes, such as abnormal insulin signaling and insulin resistance, are also pathological features of dementia.^14^ Lipids are another risk factor that deserves attention in the control of dementia.^15^

Until recently, it was believed that elevated levels of HDL cholesterol were beneficial for health. ^16^ Indeed, numerous studies demonstrated protective associations between elevated HDL cholesterol levels and reduced risk of heart disease, inflammatory conditions, and even cognitive decline.^17^ The protective effects of HDL may be attributed to its antioxidant and anti-inflammatory properties, as well as its ability to remove excess ’bad’ cholesterol.^18^ Based on several such studies, it has even suggested that increasing HDL levels or restoring its functions could be explored as a therapeutic option to combat inflammation and AD.^19,20^

Emerging evidence, however, has now challenged this established understanding.^21^ Despite these findings, there is a lack of evidence regarding the connections between dementia risk factors in the brain and in the blood. The combination of risk factors might drive individual differences in dementia progression.^10^ Therefore, examining the combination of hypometabolism risk factors, such as blood glucose and HDL-C levels, may provide more insights into individual differences in dementia progression.

The presence of the *APOE4* allele significantly increases the risk of developing AD.^22^ Despite numerous research studies, many aspects of the role of *APOE4* in AD remain unclear, including its interaction with dementia risk factors.^23^ Due to these reasons, we also sought to explore how the effects were modified when these risk factors were present in carriers of *APOE4*.

## Methods

### Data Source

We conducted this analysis using the Alzheimer’s Disease Neuroimaging Initiative (ADNI) database. The ADNI project was started in 2004 with Michael Weiner as the chief investigator. This project is a part of public-private collaboration that includes major institutions across North America. ADNI encourages the cost-free sharing of deep genotyped and phenotyped datasets with interested researchers worldwide. ADNI collects biomarkers, brain scans, clinical data, and cognitive assessments from volunteer participants based on a preset inclusion criteria. ADNI seeks to improve clinical prediction of AD and its treatment by leveraging the wide variety of available longitudinal neuroimages, biomarkers, and cognitive scales.

### Details of FDG PET Imaging

FDG PET imaging was performed on a subset of participants based on a standard protocol. Data gathered within ADNI are allocated to different cores, each comprising experts specializing in corresponding domains, tasked with efficiently managing the data.^24^ There are two imaging cores, with PET imaging falling under the jurisdiction of Banner Institute, which specifies parameters to maintain optimal imaging quality. To ensure comparability and quality across various scanners, a 3D correction is applied to the acquired images. For more information on PET measurements in the ADNI study, refer to Mueller et al.’s publication.^8^

### Generation and Interpretation of Hypometabolic Convergence Index Scores

We investigated the longitudinal changes in hypometabolic convergence index (HCI) as the study outcome. HCI values were accessed from the BAIPETNMRCFDG dataset (https://adni.bitbucket.io/reference/baipetnmrc.html). The HCI was generated from comprehensive whole-brain image analysis rather than regional examination of FDG PET brain images, and summarizes the extent of brain hypometabolism in the form of z-scores.^25^ These scores were obtained through voxel-wise analysis of the images using Statistical Parametric Mapping (SPM) software.^26^ An increasing HCI was interpreted as indicative of greater brain hypometabolism.^25^

### Covariate Extraction, Measurements, and Data linkage

We extracted glucose and lipid biomarkers from the ADNINIGHTINGALELONG dataset. In ADNI, *APOE4* was measured using DNA extracted by Cogenics from a 3 mL aliquot of EDTA blood collected during participant screening visits.^27^ Only rows containing values for all the variables were further considered for the analysis. Information on smoking and alcohol use was obtained from the medical history file. Data regarding systolic and diastolic blood pressure, sex, marital status, race, and education were derived from the vitals and demographics datasets. Age was calculated as the difference in years between the examination date and the date of birth for each participant over time. Diabetes medications were extracted from the medication data using the Anatomical Therapeutic Classification (ATC) codes (https://www.who.int/tools/atc-ddd-toolkit/atc-classification). When applicable, we used the visit code and distinct participant identifier to link datasets. Once linked, a participant was considered to be taking diabetes medication for all subsequent data following the initial prescription. We utilized the tidyr package’s *fill* function with the ‘direction=down’ option to propagate the diabetes medication use labels.

### Statistical Analysis

We performed data analysis and visualizations using the R programming language (R version 4.3.2). ^28^ At first, variable summaries measured at baseline were computed. For this, continuous variables were presented as mean with standard deviation. Categorical variables were summarized as frequencies and percentages. To depict correlations between continuous covariates at the baseline, we employed a correlation heatmap. To assess longitudinal variations in plasma glucose and HDL-C levels, we pooled all observations from all visits and calculated the coefficient of variation percentage (CV%). CV% was computed by dividing the standard deviation by the mean and multiplying by 100. We generated a scatter plot of the CV% for glucose and HDL-C distributions to assess their relationship. Additionally, we plotted the CV% for these measures stratified by *APOE4* allele status and determined if the differences were statistically significant using the Kruskal-Wallis test.

To examine the relationship between blood glucose, HDL-C levels, *APOE4* status, and the progression of brain hypometabolism, and to account for the correlated data structure, we conducted a linear mixed model analysis using the lme4 package.^29^ Variations between participants and visits were accounted for by specifying random intercepts and varying slopes in the random effects part of the model. We analysed multiple models with different combinations of terms and then selected the best model with the lowest Akaike Information Criterion (AIC) values. This model was deemed the model with the minimum set of variables that best explains the data. Model comparisons were performed using the anova built-in function. For unbiased regression estimates in the optimal model, we employed restricted maximum likelihood estimation. The Nelder-Mead optimizer was used to ensure model convergence.

To analyze the conditional and marginal variable contributions of the covariates in the parsimonious mixed model, we utilized the hierarchical partitioning method.^30^ This approach helps to understand how *APOE4*, HDL-C, and glucose modify the HCI. We computed marginal means and corresponding 95% confidence intervals (CI) to quantify the average change in HCI for these variables.

We also checked the functional relationship between glucose and HDL-C with HCI using generalized additive mixed effect models (GAMM). The advantage of GAMM is that it is able to incorporate the benefits of generalized additive models, i.e., nonlinear effects modeling, while also accounting for correlations due to repeated measures.^31^ The AIC from both linear and nonlinear models were compared to determine the most suitable functional relationship between the variables. A two-tailed p-value <0.05 was considered statistically significant. Additionally, we quantified and visualized individual heterogeneity for the random effects terms specified in the optimal mixed effects model. Lastly, to assess the HCI reduction associated with varying values of glucose and HDL-C, we generated a partial effect plot.

## Results

### Sample description

Data on 336 individuals (781 observations) were available for analysis after data linkage, with 22.62% of participants diagnosed with AD (Supplementary Figure 1). Table 1 describes the baseline demographic and clinical characteristics of the participants in this study. The participants were, on average, 75.43 years old and had 15.6 years of education. The majority were male (63.98%) and White (93.45%). Over a third (38%) had a smoking history, and 97.6% reported being ever married. The mean HbA1C and HDL-C levels were 5.42 and 1.52, respectively. The mean systolic blood pressure (SBP) was relatively high at 135 mm Hg, whereas the mean diastolic blood pressure (DBP) was below the normal reference level at 73.77 mm Hg. Almost half (48.51%) were carriers of the *APOE4* allele. Figure 1 shows the distribution of longitudinal CV% for glucose and HDL-C, as well as their relationship. In Figure 2, the distributions of longitudinal CV% for glucose and HDL-C, stratified by *APOE4* allele status, are shown. Non-linear modeling (spline fit) of the relationship between CV% for glucose and HDL-C supports a nonlinear relationship. P-values from the Kruskal-Wallis test to see the influence of *APOE4* alleles on the CV% for glucose and HDL-C were non-significant (0.78 and 0.56, respectively). Brain hypometabolism, measured by HCI, was higher in this sample, with a mean of 15.06 (range 4.34 -47.40). Initially, only 2 participants (0.60%) were using anti-diabetes medication, but by the end of the follow-up, this number had risen to 10 (2.97%). As regards to the baseline correlations, *APOE4* was positively correlated to HCI (Supplementary Figure 2). *APOE4* had no strong correlations with either lipid subgroups and blood glucose. In those with AD, blood glucose and HDL-C modelled using splines seems to favor a more non-linear relationship in comparison to those not diagnosed with AD (Supplementary Figures 3-4).

**Figure 1.**
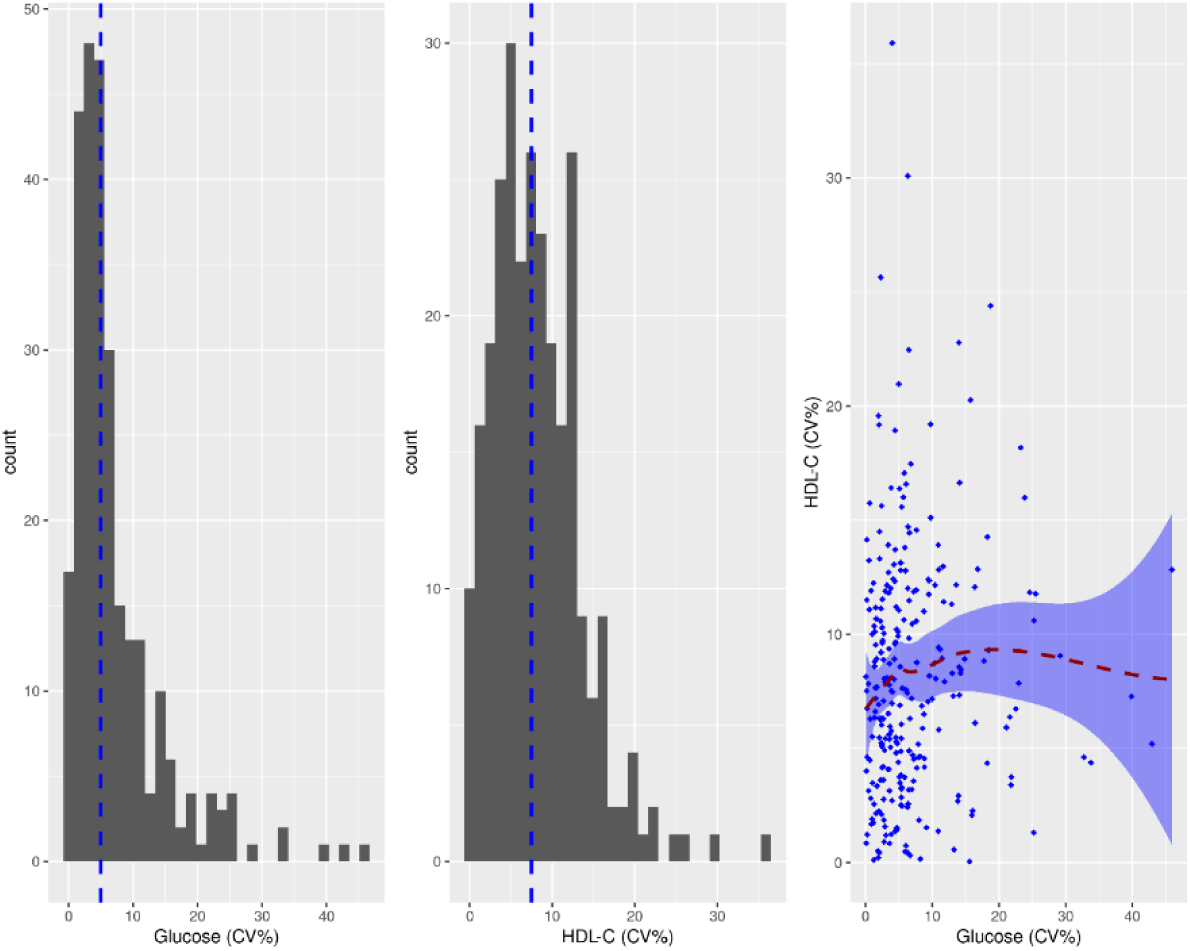
Histogram of longitudinal CV% for glucose and HDL-C, and a scatterplot with smoothed regression line showing their relationship

**Figure 2.**
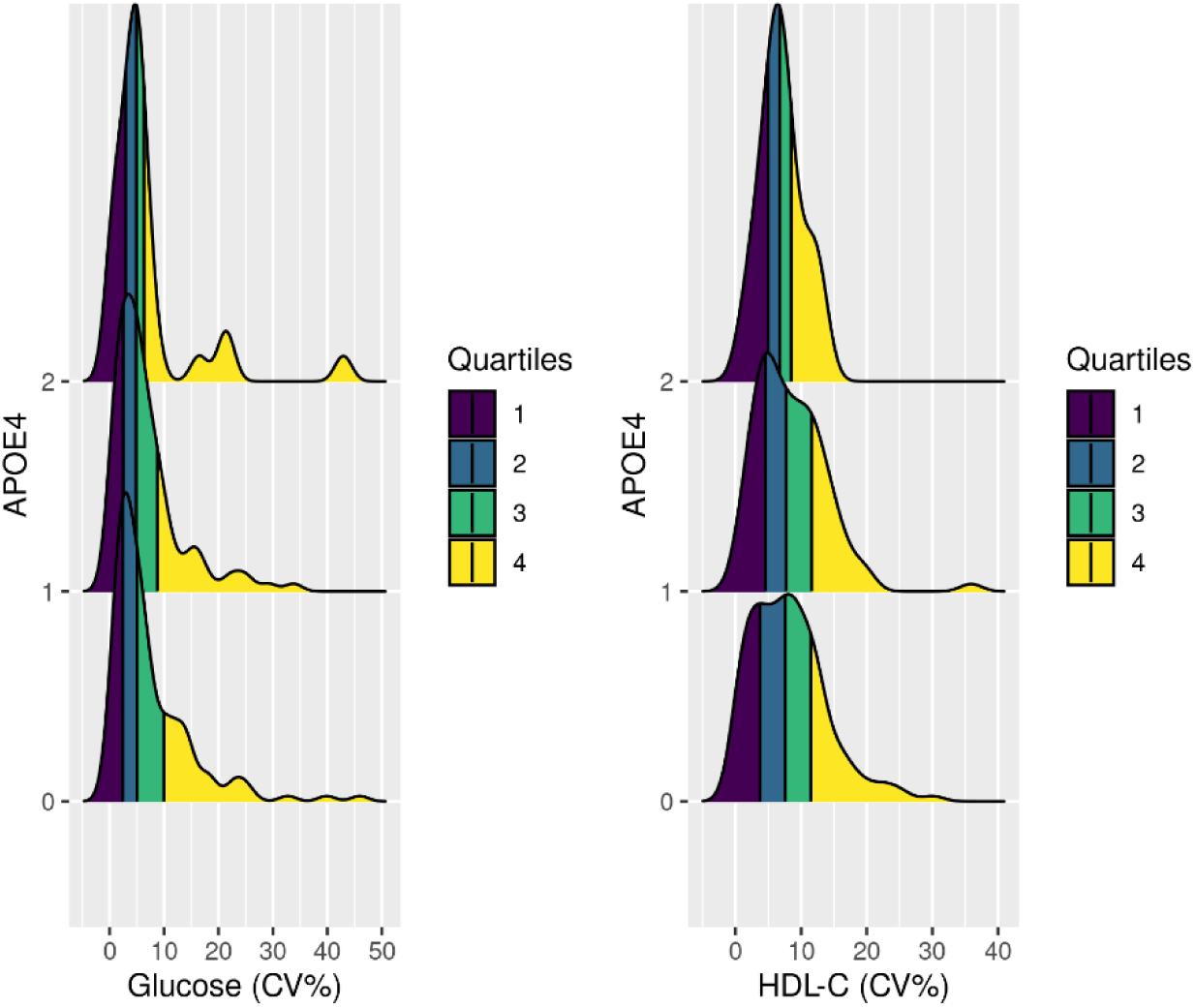
Distribution of longitudinal CV% for glucose and HDL-C stratified by *APOE4* allele status

**Table 1.**
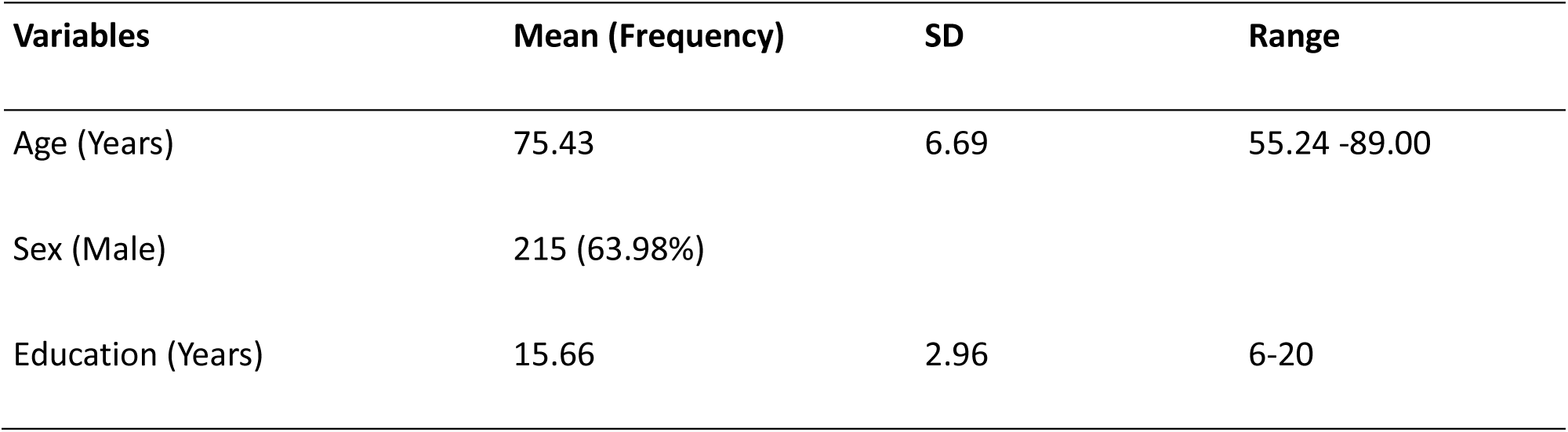
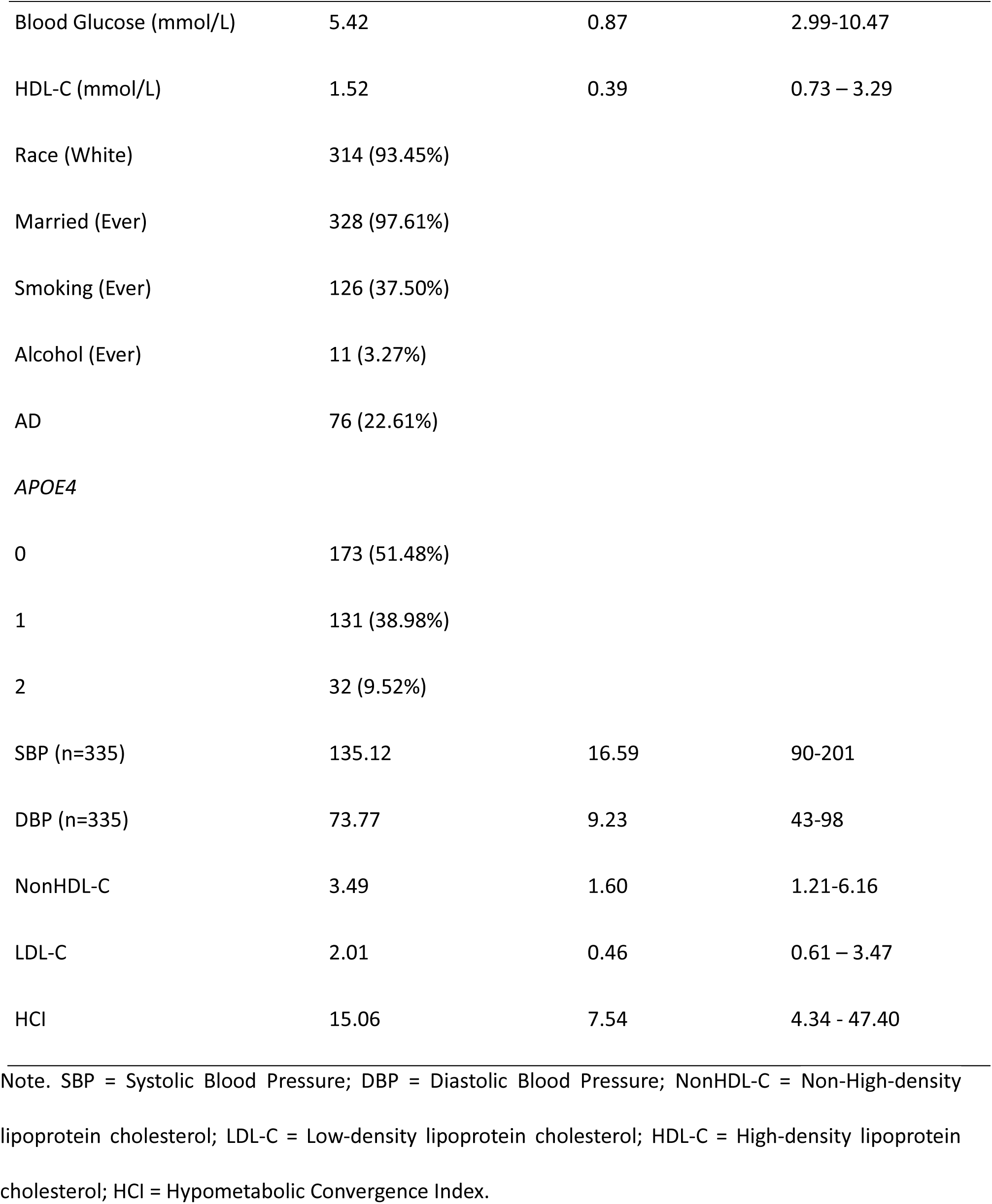
Characteristics of participants measured at baseline (781 observations, n=336)

| Variables | Mean (Frequency) | SD | Range |
| --- | --- | --- | --- |
| Age (Years) | 75.43 | 6.69 | 55.24 -89.00 |
| Sex (Male) | 215 (63.98%) |  |  |
| Education (Years) | 15.66 | 2.96 | 6-20 |
| Blood Glucose (mmol/L) | 5.42 | 0.87 | 2.99-10.47 |
| HDL-C (mmol/L) | 1.52 | 0.39 | 0.73 – 3.29 |
| Race (White) | 314 (93.45%) |  |  |
| Married (Ever) | 328 (97.61%) |  |  |
| Smoking (Ever) | 126 (37.50%) |  |  |
| Alcohol (Ever) | 11 (3.27%) |  |  |
| AD | 76 (22.61%) |  |  |
| *APOE4* |  |  |  |
| 0 | 173 (51.48%) |  |  |
| 1 | 131 (38.98%) |  |  |
| 2 | 32 (9.52%) |  |  |
| SBP (n=335) | 135.12 | 16.59 | 90-201 |
| DBP (n=335) | 73.77 | 9.23 | 43-98 |
| NonHDL-C | 3.49 | 1.60 | 1.21-6.16 |
| LDL-C | 2.01 | 0.46 | 0.61 – 3.47 |
| HCI | 15.06 | 7.54 | 4.34 - 47.40 |

Note. SBP = Systolic Blood Pressure; DBP = Diastolic Blood Pressure; NonHDL-C = Non-High-density lipoprotein cholesterol; LDL-C = Low-density lipoprotein cholesterol; HDL-C = High-density lipoprotein cholesterol; HCI = Hypometabolic Convergence Index.

### Effect of Elevated Blood Glucose and HDL-C Variability on Brain Hypometabolism

As shown in the Supplementary Table 1, Model 2 was selected for detailed analysis. Table 2 shows the adjusted regression estimates from the optimal linear mixed model. More number of clinical visits (β=1.67, 95% CI: 1.37, 1.98, p<0.001), and *APOE4* carrier status were the strongest predictors for the decline in brain metabolism in ascending order respectively (Supplementary Figure 5). The contributions of these variables to variance from fixed effects in the best model were also the highest. Over time, an increase in plasma glucose was significantly associated with an increased area of brain hypometabolism (β=0.33, p<0.05). None of the cholesterol markers except HDL-C were statistically significant in the multivariate analysis (β=1.22, p<0.05). Additionally, age, sex, smoking, blood pressure, and race were not significant predictors. The percentage variance of the fixed effects (marginal R-squared) was estimated at 4.5%. Supplementary Figure 6 elucidates the random effects represented by the clinical visits and individual variability. Variance, combining both the fixed and random effects (conditional R-squared), accounted for 95.2% of the model variance.

**Table 2.**
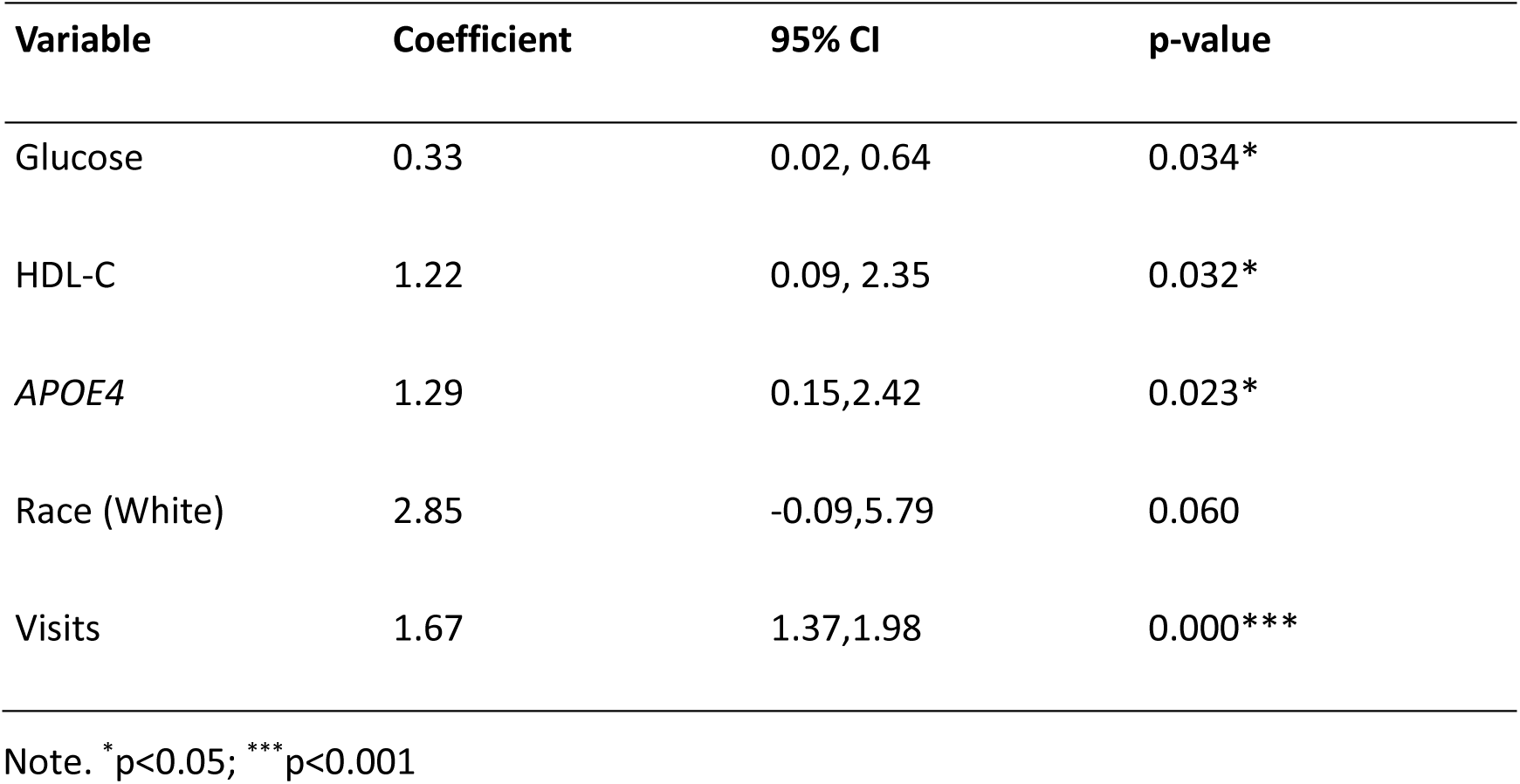
Adjusted model estimates corresponding to the optimal linear mixed model for the HCI trend outcome.

| Variable | Coefficient | 95% CI | p-value |
| --- | --- | --- | --- |
| Glucose | 0.33 | 0.02, 0.64 | 0.034* |
| HDL-C | 1.22 | 0.09, 2.35 | 0.032* |
| *APOE4* | 1.29 | 0.15, 2.42 | 0.023* |
| Race (White) | 2.85 | -0.09, 5.79 | 0.060 |
| Visits | 1.67 | 1.37, 1.98 | 0.000*** |

Note. \* $p<0.05$ ; \*\*\* $p<0.001$

### Functional Relationship Between Glycemic Variability, HDL-C Levels, and Their Interaction with *APOE4* on Brain Hypometabolism

A comparison of effects plots (Supplementary Figure 7, Figure 3, and Figure 4) generated from linear and additive mixed models (GAMM) indicates that glucose has a linear relationship with HCI. Conversely, HDL-C showed a nonlinear relationship, which was preferred over the linear model. Additive mixed models work similarly to linear mixed models, with the difference being that they allow for modeling complex non-linear relationships of variables by fitting smooth functions.^31^ Partial effects here refer to the mean effect in HCI due to the change in exposures while keeping the effects of other covariates held constant in the model. The dose-response relationship between HDL-C and HCI was observed to decrease slightly, stabilize and then decline after 2 mmol/L for higher values. According to Figure 4, elevated HDL-C reduces brain metabolism even when glucose levels are optimal. Hypometabolism increases with each unit rise in glucose, particularly above 7.5 mmol/L.

**Figure 3.**
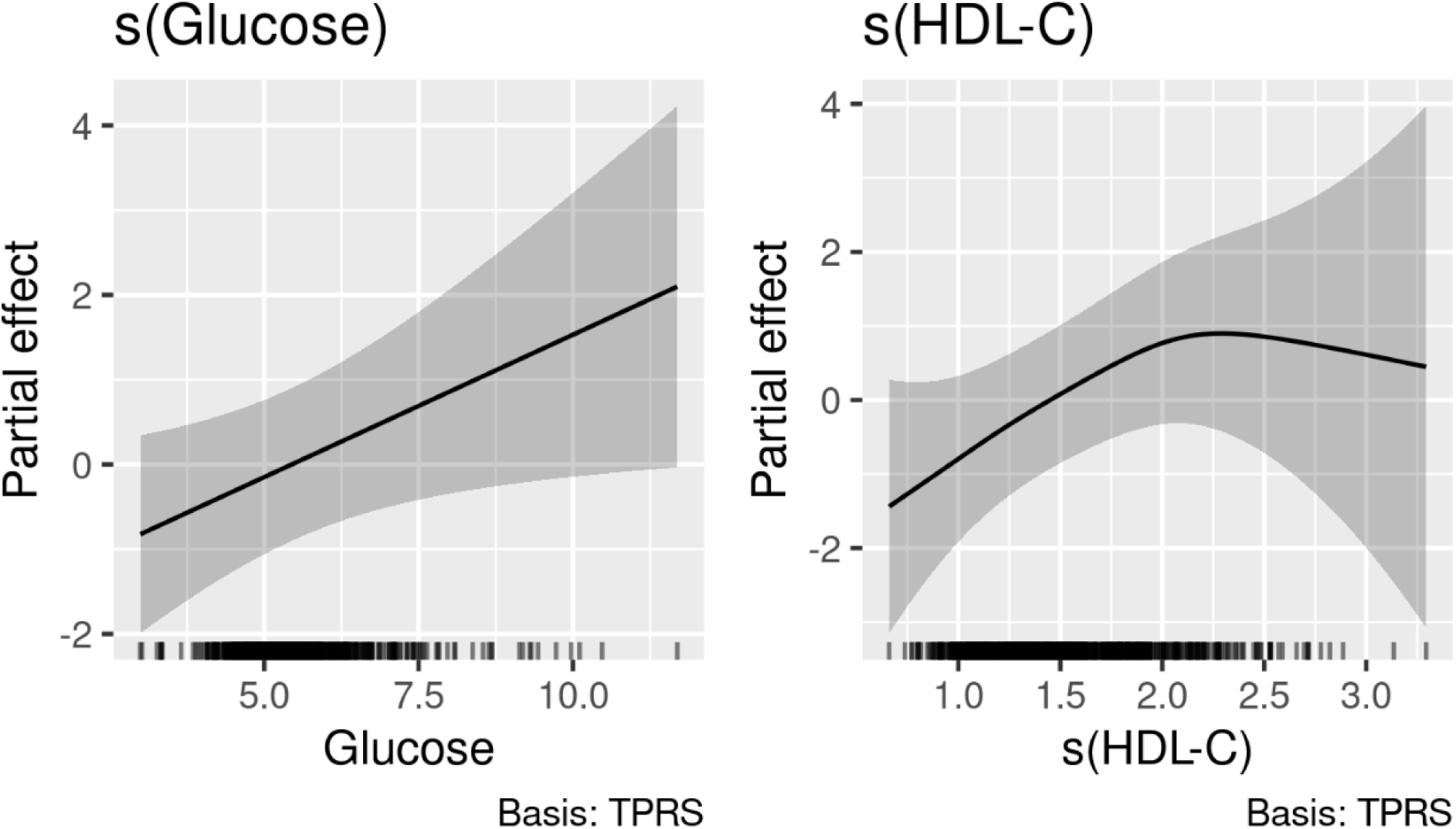
Partial effects of Glucose and HDL-C levels on the HCI from the GAMM model. Note. TPRS (thin plate regression splines) are basis functions that allow for model fitting of local segments of the exposure-outcome relationship, which are then connected to provide a complete picture of the overall relationship. Thin plate splines automatically determine the location and number of knots based on changes in the values of the covariate.

**Figure 4.**
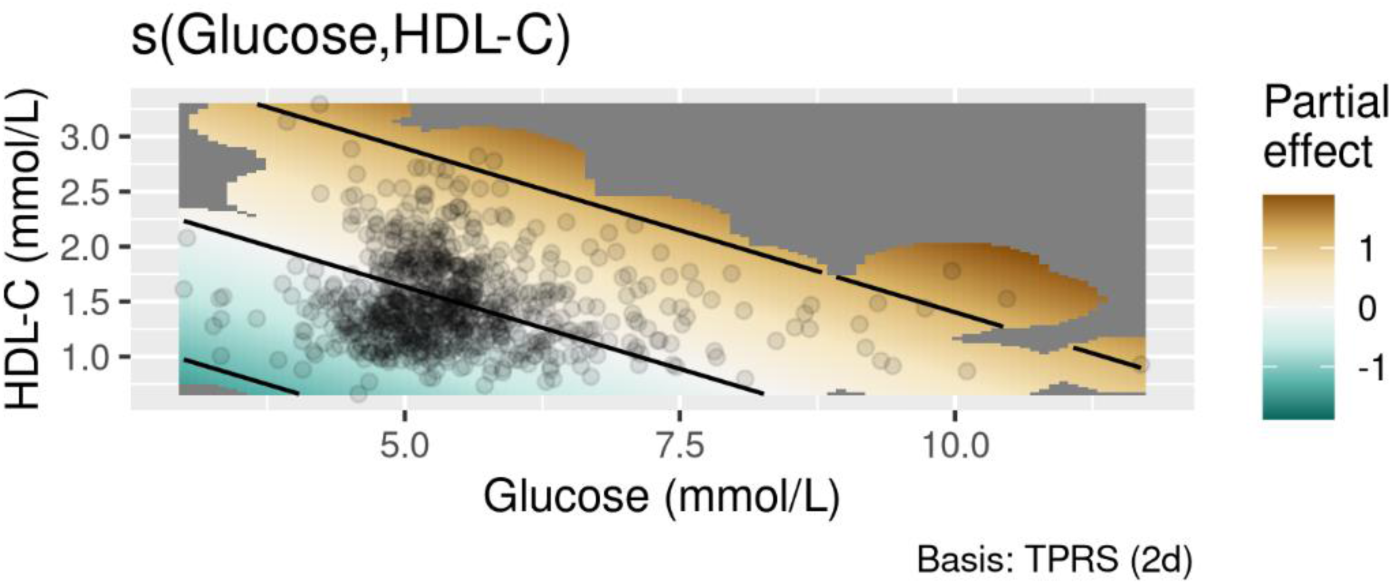
The partial effect plot illustrates the combined effects of HDL-C and glucose levels on HCI. The variable relationships are shown as a smoothed relationship as a function of these variables and are estimated from the additive mixed model. In the plot, the black lines and black dots represent the contour lines and observed data points, respectively. A change in color from green to orange reflects how the relationship changes, i.e., from a negative to a positive partial effect. A partial effect of -1 means that a one-unit increase in the predictor variable is associated with a one-unit decrease in the outcome variable, and vice versa. It is evident that the partial effects vary at different levels of the predictor variables.

At this threshold, risk is evident even for low HDL-C values. Hence, it could be deduced that elevated glucose values are an important risk factor for brain hypometabolism, and that its effects with HDL-C on brain metabolism are non-linear. Supplementary Figure 8 complements this finding, showing substantial differences in predicted HCI across stratified HDL-C categories of low, intermediate, and high levels. Similar trends were evident in the interaction effects of blood glucose levels, HDL-C, and the *APOE4* allele on HCI (Figure 5). The plot shows a dramatically pronounced decline in brain metabolism for increased blood glucose levels and HDL-C in *APOE4* homozygous carriers compared to non-carriers and heterozygous carriers. In addition, for better interpretation of the slopes, the marginal means for the HCI, computed for the different combinations of these risk factors, are shown in Supplementary Table 2. Compared to non-carriers with low HDL-C and high glucose, there was a more than 10-point increase in predicted mean HCI for double *APOE4* carriers with high HDL-C.

**Figure 5.**
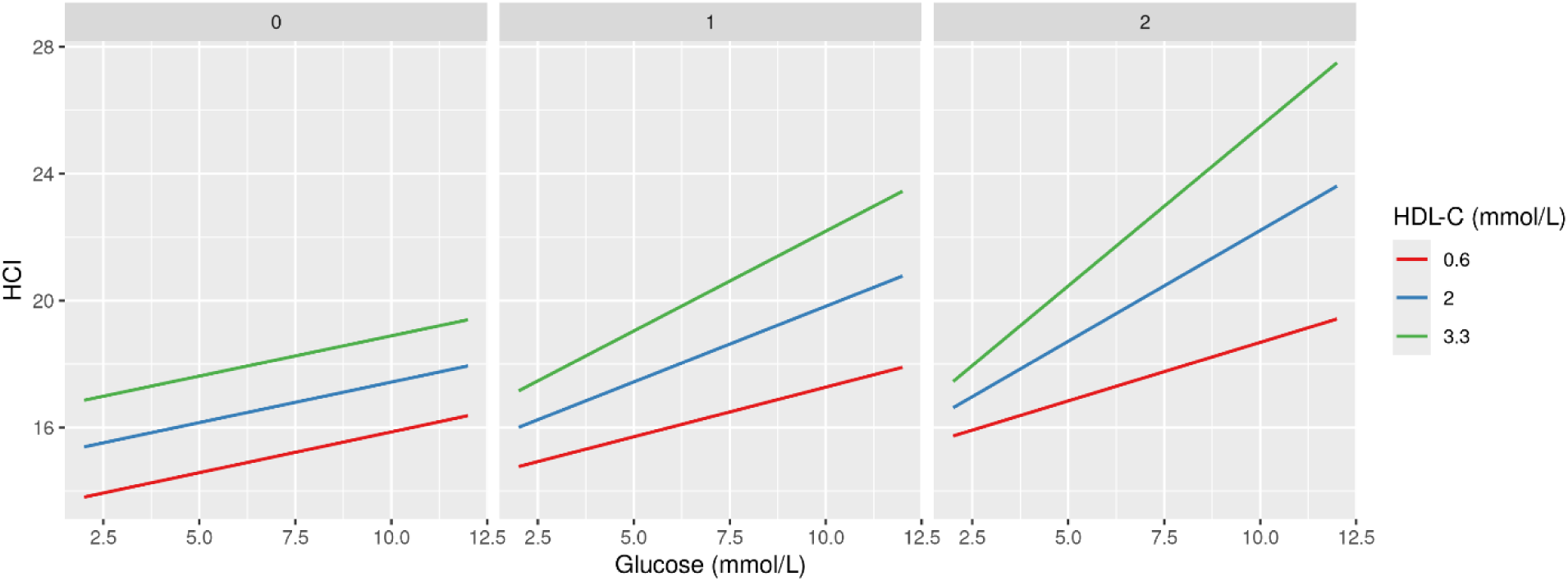
Plot showing the synergistic effects of blood glucose levels, HDL-C, and *APOE4* allele on HCI. The predictions were generated from a model containing interaction terms for blood glucose levels, HDL-C, and *APOE4*. Each window corresponds to the effects of the *APOE4* allele (0, 1, 2) and is shown specifically for HDL-C categories (low, high, and intermediate, arbitrarily selected based on the data)

### Comparison of Subgroup Analysis to Identify Heterogeneity in Effects

Given that the visualizations indicated a possible difference in the effects of the exposures in AD and non-AD individuals, we prepared the data accordingly. Upon further investigation, we noted that the statistical associations were limited to the non-AD subgroup (Supplementary Tables 3-4). Remarkably, the strength of the associations for White race, HDL-C, and *APOE4* with HCI was much stronger in the stratified analysis for this group. Overall, the effect size for glucose did not differ for the non-AD individuals compared to the main analysis, whereas the clinical visits exhibited a diluted effect. For the AD group, both *APOE4* and visits were still significantly associated with HCI. While the impact of visits was considerably stronger than in the aggregated model, the effects of *APOE4* were strongly reversed, indicating a protective association.

## Discussion

In this study of older adults, plasma glucose and HDL-C variability were significantly associated with a reduction in brain metabolism, but only in individuals without AD. Importantly, these effects were independent of *APOE4* and common confounders such as age, sex, and other lipids profiles. We also found that the relationship of plasma glucose and HDL-C with brain metabolism operates independently of each other. *APOE4* status and measurement visit were the strongest predictors of brain metabolism at the population level. This is not surprising, as we demonstrated in our previous work using the ADNI data that measurement visits provide more information than age alone.^32^ Measurement visits may indirectly reflect patient characteristics, response, and observation time.^33^

A hyperglycemic milieu can cause widespread systemic effects, thereby modulating physiological responses.^34^ It is important to note that the diabetes burden was quite low in our data. However, diabetes is not an absolute requirement for the glucose to impact brain. Our findings are consistent with a previous study which demonstrated that even midlife increase in glucose can accelerate dementia.^35^ Although hyperglycemia contributes to the development of dementia, its role in certain types of dementia such as in AD remains to be established.^36^ In our case, one possible reason could be that participants had AD at baseline and, therefore, already had much lower brain metabolism. Therefore, it is much less likely that glucose and HDL-C variations would have any impact. Also, *APOE4* is no more a risk factor since they had already developed AD. Hyperglycemia-induced physiological changes are quite complex and usually do not conform to the normal dose-response framework.^37^ However, in our study, an increase in blood glucose levels was linearly related to a decline in brain metabolism. This contradicted a previous study in which cognitive decline was observed with both high and low blood sugar levels, and it worsened with age.^38^ Nevertheless, high blood sugar-driven outcomes at the individual level are highly heterogeneous and also depend on the combination of other risk factors and genes.^39,40^

In our data, elevated HDL-C was stronger risk factor for brain hypometabolism than plasma glucose levels. The role of lipids in dementia remains controversial. A meta-analysis of multiple studies published on lipid subgroups shows that LDL-C could be a likely candidate risk factor for dementia, but there was no evidence of involvement of HDL-C or other lipids.^41^ On the other hand, it is worth noting that high HDL-C levels could negatively impact health and survival, including increased all-cause and cardiac-related mortality. However, individuals in the mid-range levels of HDL-C seem to be protected.^42^ This was observed in the ’U-shaped’ relationship between HDL-C levels and cognitive outcomes, with individuals having HDL-C values above 2.50 mmol/L experiencing more than a two-fold increased risk of poor cognitive outcomes.^43^ Such a relationship was also reported in a large-scale survival analysis of health insurance data exploring dementia outcome. Similar to our study, LDL-C in this study did not influence dementia risk, except for a minor increase in risk observed among statin users.^44^

A study published in Lancet, that investigated whether HDL-C is a risk factor for incident dementia reported a risk above 3.3 mmol/L. ^45^ Consistent with the previous findings, individuals aged above 75 years with high HDL-C were at substantial risk for dementia. In stark contrast, lower HDL-C values were shown to be protective against dementia.^45^ This was in line with our finding. Based on the partial effects plots in our study, HDL-C associated risk appeared to diminish beyond 2.25 mmol/L. However, these findings require further validation, as there were only a few HDL-C values above this threshold in the data. It might be that the HDL-C effects seen were due to the presence of AD, co-morbidities, or other age-related factors. In such scenarios, high HDL would be merely reflective of these conditions rather than providing any real health benefits.^20^

*APOE4* carriage was associated with a greater metabolic decline compared to non-carriers for the concomitant values of glucose and HDL-C. Notably, this interaction was particularly strong in individuals with HDL-C levels above 2 mmol/L. It is intriguing that the direction of the predicted slope for brain hypometabolism with glucose elevation was similar at lower HDL values across all *APOE4* isoforms, with carriers experiencing a slightly higher metabolic decline. The observed interaction effects appear plausible as *APOE4* have a direct link with cholesterol metabolism and lower HDL synthesis.^46–48^ In addition it has been observed that mice with *APOE4* risk alleles exhibit a poor response to glucose spikes and inadequate insulin production.^49^ Individuals with *APOE4* risk alleles and high blood glucose were more likely to experience greater risk for severe dementia and features of AD in late life.^35^ First of all, carrying two *APOE4* alleles is now considered a genetic form of AD by itself.^50^ Our finding regarding the synergistic effect modulated by *APOE4* double carriers is in firm agreement, showing a vastly different pattern of hypometabolism compared to its other isoforms. Therefore, the effects in those without AD might actually be congruent with preclinical AD.^51^ This should be kept in mind, especially in the context that the diagnostic criteria for AD are still evolving.^51^ *APOE4* carrier status favors AD and dementia mainly through promoting amyloid beta, increased phosphorylated tau, and contributing to neurodegeneration.^27,52,53^ It is also recognized that *APOE4* variation can negatively impact mitochondrial respiration and energy production, consequently leading to brain hypometabolism.^54^ Other potential pathways include neuroinflammation, blood-brain barrier dysfunction, gliosis, brain structural and functional changes, demyelination and impaired clearance of toxic substances.^55–60^

In relevance to our work, it has already been demonstrated that *APOE4* variation adversely impacts the ability of HDL-C to effectively sequester cholesterol by modifying HDL-C structurally. Not only does the concentration of HDL matter, but also its size. For instance, individuals with AD and dementia tend to have relatively smaller HDL particle sizes.^61^ We presume that *APOE4*-induced changes in the brain may make it more vulnerable to the negative physiological effects of elevated glucose and other risk factors. As corroborating evidence, we found no indication that longitudinal variability in glucose and HDL-C can be attributed to *APOE4* alleles.

This study has several strengths including the availability of serial participant data and a well-characterized cohort. Our study is among the first to illuminate the joint contributions of glucose, HDL-C, and *APOE4* allele variations on brain hypometabolism. We were able to clearly demonstrate the change in relative importance of these major risk factors through visualizations. This approach provides a more comprehensive understanding of the potential pathophysiology, which may not be fully revealed when examining these factors individually.

Our study had limitations. Blood glucose and HDL-C collection was not timed according to disease pathology. The study did not account for comorbidity status or medication use as covariates, with the exception of blood pressure, and diabetes. We have not specifically investigated the possibility of sexual dimorphism in our results, nor have we considered the involvement of potential mechanisms such as amyloid pathways or other inflammatory markers. Another constraint was the limited representation of non-white samples, which restricted our ability to compare and explore differences across racial groups. This, combined with the smaller sample size and selective recruitment in the cohort, may further limit the generalizability of the findings. Lastly, there could be unmeasured confounding. Therefore, we recommend replicating our findings in large cohorts and across multiple ethnicities to ascertain the benefits of glucose and HDL reduction in diverse populations. Further studies may be conducted using genetic variants that influence these exposures to gather causal evidence.

## Conclusion

High blood glucose levels facilitated progression of cerebral hypometabolism in ADNI participants. The negative impact of blood glucose on brain hypometabolism was aggravated by elevated HDL-C levels and *APOE4* carrying status. High blood glucose levels may reflect systemic insulin resistance, which, in turn, might impair brain glucose uptake, resulting in brain hypometabolism. While further validation is warranted, controlling for plasma glucose/insulin resistance and HDL-C levels in *APOE4* carriers may attenuate the decline in brain metabolism, potentially delaying dementia clinical onset.

## Abbreviations

AD: Alzheimer’s Disease
ADNI: Alzheimer’s Disease Neuroimaging Initiative
ATC: Anatomical Therapeutic Classification
CV%: Coefficient of Variation Percentage
DBP: Diastolic Blood Pressure (DBP)
FDG: Fluorodeoxyglucose-18 (FDG)
GAMM: Generalized Additive Mixed Effect Models
HCI: Hypometabolic Convergence Index
HDL-C: High-density lipoprotein cholesterol
LDL-C: Low-density lipoprotein cholesterol
NonHDL-C: Non-High-density lipoprotein cholesterol
PET: Positron Emission Tomography
SBP: Systolic Blood Pressure (SBP)
SPM: Statistical Parametric Mapping
TPRS: Thin Plate Regression Splines

## Data Availability

All data produced are available online at https://adni.loni.usc.edu/

## Acknowledgements

The authors would like to thank the ADNI team. Data collection and sharing for this project was funded by the Alzheimer’s Disease Neuroimaging Initiative (ADNI) (National Institutes of Health Grant U01 AG024904) and DOD ADNI (Department of Defense award number W81XWH-12-2-0012). ADNI is funded by the National Institute on Aging, the National Institute of Biomedical Imaging and Bioengineering, and through generous contributions from the following: AbbVie, Alzheimer’s Association; Alzheimer’s Drug Discovery Foundation; Araclon Biotech; BioClinica, Inc.; Biogen; Bristol-Myers Squibb Company; CereSpir, Inc.; Cogstate; Eisai Inc.; Elan Pharmaceuticals, Inc.; Eli Lilly and Company; EuroImmun; F. Hoffmann-La Roche Ltd and its affiliated company Genentech, Inc.; Fujirebio; GE Healthcare; IXICO Ltd.;Janssen Alzheimer Immunotherapy Research & Development, LLC.; Johnson & Johnson Pharmaceutical Research & Development LLC.; Lumosity; Lundbeck; Merck & Co., Inc.;Meso Scale Diagnostics, LLC.; NeuroRx Research; Neurotrack Technologies; Novartis Pharmaceuticals Corporation; Pfizer Inc.; Piramal Imaging; Servier; Takeda Pharmaceutical Company; and Transition Therapeutics. The Canadian Institutes of Health Research is providing funds to support ADNI clinical sites in Canada. Private sector contributions are facilitated by the Foundation for the National Institutes of Health (www.fnih.org). The grantee organization is the Northern California Institute for Research and Education, and the study is coordinated by the Alzheimer’s Therapeutic Research Institute at the University of Southern California. ADNI data are disseminated by the Laboratory for Neuro Imaging at the University of Southern California.

## Funding

This research was funded by the National Institute on Aging of the National Institutes of Health under award numbers R01AG070487 and R01AG062623. The content is solely the responsibility of the authors and does not necessarily represent the official views of the National Institutes of Health.

## Contributions

KA, SU, and ALR were involved in conceptualization, design and wrote the paper. OB, ALR, MD conducted data curation and formal analysis. KA, and AIY provided guidance on data analysis, interpreted the findings, and critically revised the manuscript. All authors have read and agreed to the published version of the manuscript.

## Ethics declarations

The studies involving human subjects were approved by the Duke University Health System Institutional Review Board (Protocols Pro00105166 and Pro00105389). This publication includes only secondary analyses of existing data available from ADNI, and does not include identifiable human data. Written informed consent for ADNI participants was obtained by the ADNI in accordance with the local legislation and ADNI requirements.

## Consent for Publication

Not applicable

## Competing Interests

The authors declare no competing interests.

## Additional information

### Electronic Supplementary Material

Supplementary Material 1

## Notes

### Competing Interest Statement

The authors have declared no competing interest.

