## Supplementary Material 1 for "Elevated blood glucose levels are associated with the progression of brain hypometabolism, and HDL-C and *APOE4* add to this association"

### Supplementary Materials

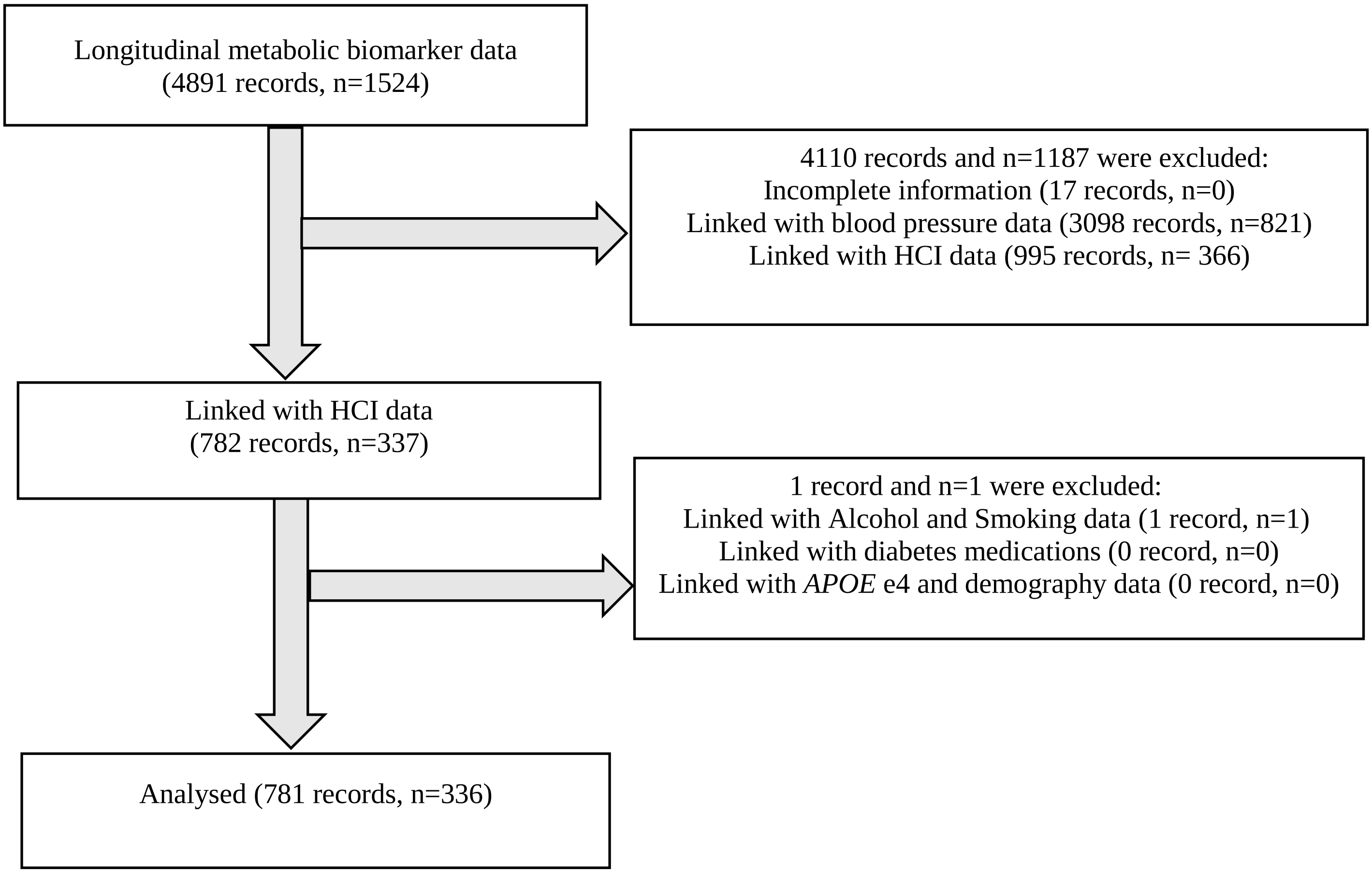

**Supplementary Figure 1.** Study flow diagram of sample selection

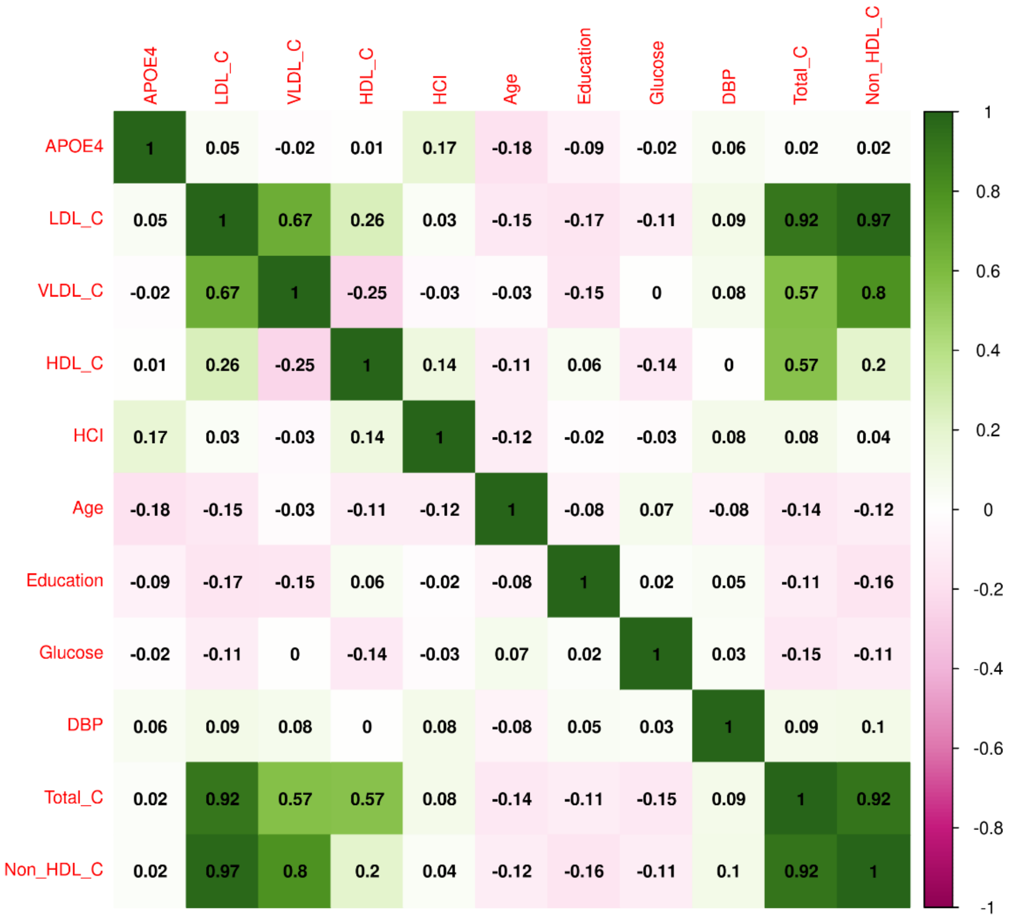

**Supplementary Figure 2.** Pearson correlation between continuous covariates at the baseline.

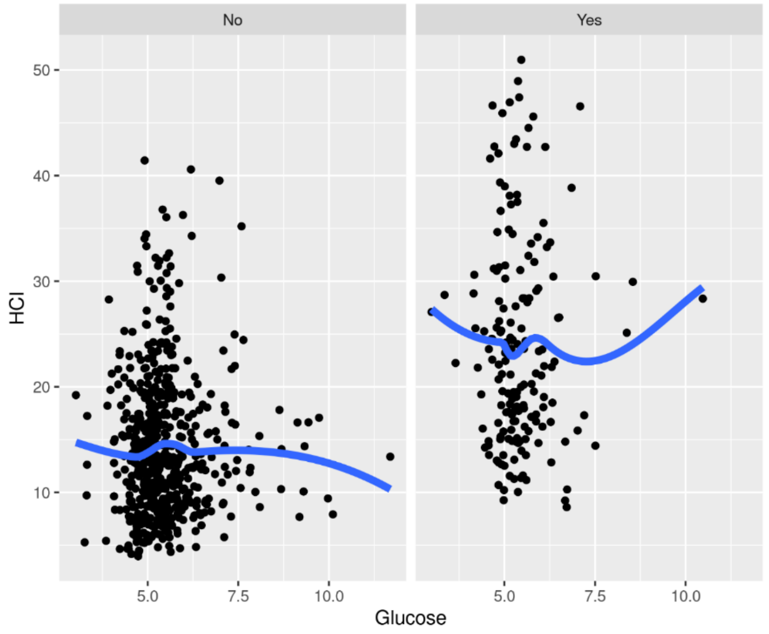

**Supplementary Figure 3.** Smoothed regression line for the relationship between plasma glucose and HCI by AD status

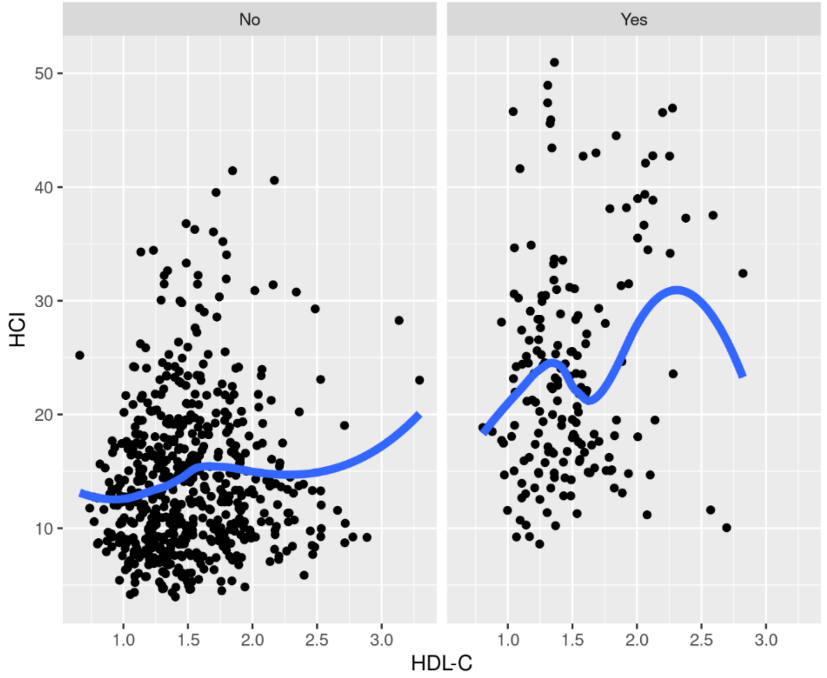

**Supplementary Figure 4**. Smoothed regression line for the relationship between plasma glucose and HDL-C by AD status

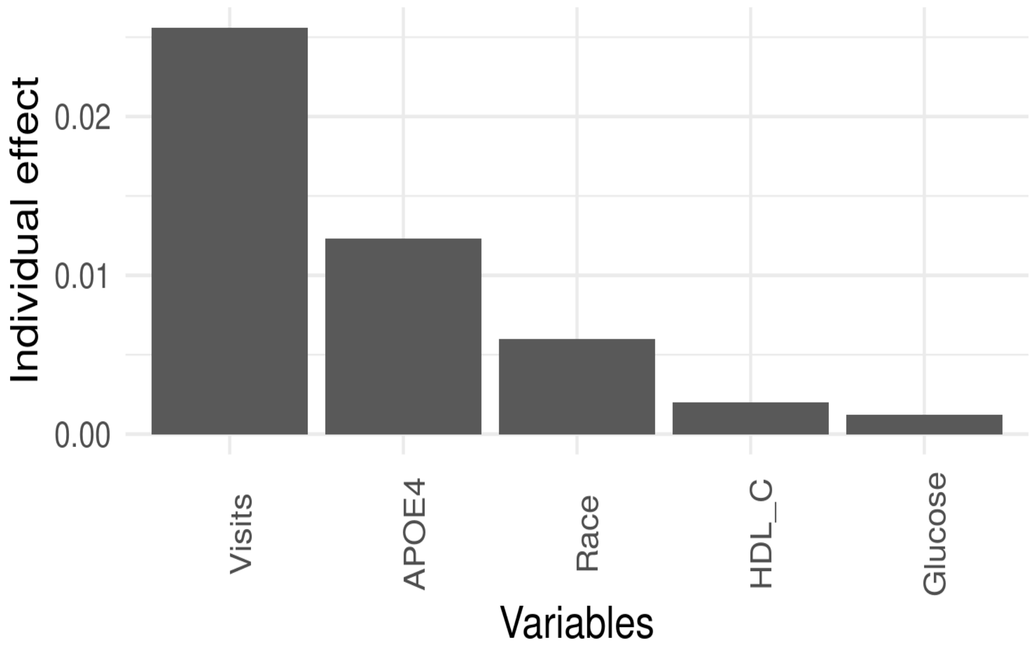

**Supplementary Figure 5.** Contribution of significant covariates to the variation accounted by fixed effects

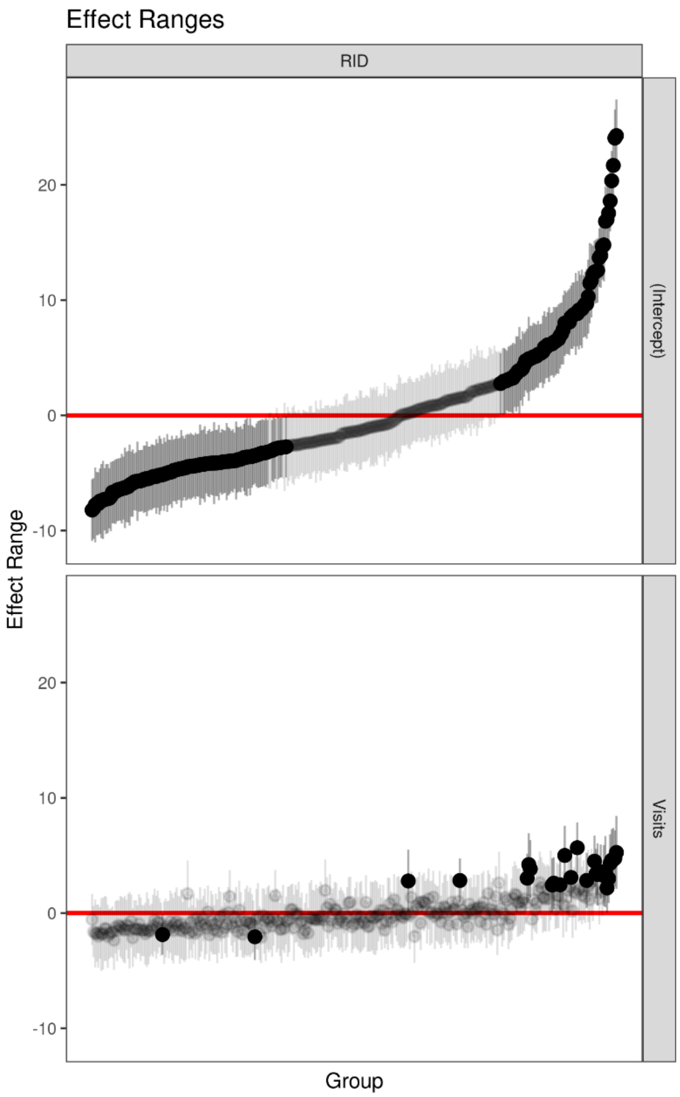

**Supplementary Figure 6**. Individual heterogeneity represented by random effects from the optimal mixed effects model: top figure - individual intercepts, and bottom figure - visits. Black dots represent values that start low or high, deviating from the average, while dim areas near the red horizontal line represent average participant values.

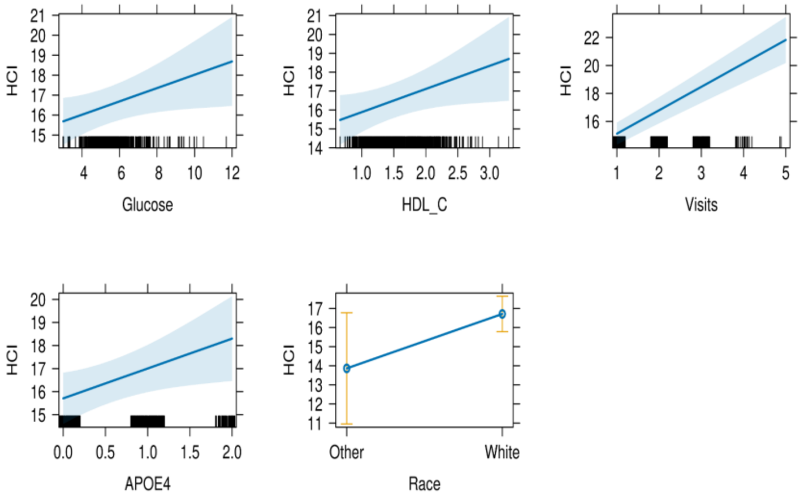

**Supplementary Figure 7.** Effect plots displaying the relation between the fixed effect predictors from the optimal linear mixed model. The black line at the bottom of the plots represents the density of observations corresponding to covariates in the dataset.

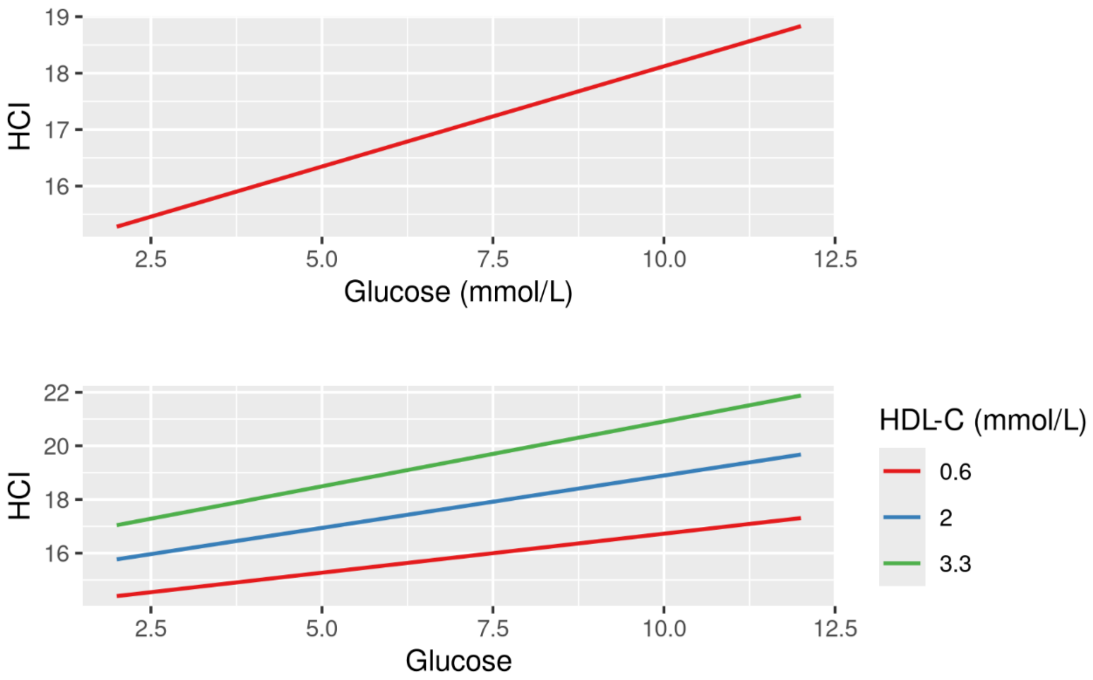

**Supplementary Figure 8.** The effect of a unit increases in plasma glucose on HCI (top), and the bottom figure illustrates the relationship of HCI with glucose levels, stratified across HDL-C categories. The HDL-C strata represent low, intermediate, and high levels, arbitrarily selected based on the data.

**Supplementary Table 1.** Model parameters from the different linear mixed models investigated as part of identifying the parsimonious model

| **Model** | **AIC** | **logLik deviance** | **p** |
|---|---|---|---|
| Model 1 | 4720.3 | -2353.1 | |
| Model 2 | 4593.0 | -2286.5 | <0.001*** |
| Model 3 | 4593.8 | -2285.9 | 0.27 |
| Model 4 | 4600.7 | -2282.3 | 0.41 |

Note. $^{*}p<0.05$; $^{***}p<0.001$

**Models:**

Model1: HCI ~ Glucose + HDL-C + Visits + *APOE4* + (1 | Participant)

Model2: HCI ~ Glucose + HDL-C + Visits + *APOE4* + Race + (1 + Visits | Participant)

Model3: Glucose + HDL-C + Visits + *APOE4* + Race + Diabetes medication+ (1 + Visits | Participant)

Model4: HCI ~ Glucose + HDL-C + Visits + *APOE4* + LDL-C + Age + Sex + Education + Smoking + Married status + SBP+ DBP + Race + (1 + Visits | Participant)

**Supplementary Table 2.** Predicted mean HCI and 95% CI for plasma glucose by *APOE4* alleles and varying HDL-C levels (low, high, and intermediate)

| Variable levels | Glucose | Predicted HCI | 95% CI |
|---|---|---|---|
| ***APOE4*: 0 HDL-C: 0.6** | 2 | 12.89 | 8.86, 16.93 |
| | 4 | 13.41 | 10.77, 16.04 |
| | 8 | 14.43 | 11.04, 17.83 |
| | 12 | 15.46 | 8.61, 22.31 |
| ***APOE4*: 0 HDL-C: 2** | 2 | 14.48 | 10.99, 17.96 |
| | 4 | 14.99 | 12.76, 17.22 |
| | 8 | 16.01 | 13.08, 18.94 |
| | 12 | 17.03 | 11.01, 23.05 |
| ***APOE4*: 0 HDL-C: 3.3** | 2 | 15.95 | 7.45, 24.44 |
| | 4 | 16.46 | 11.91, 21.00 |
| | 8 | 17.47 | 10.57, 24.37 |
| | 12 | 18.49 | 2.74, 34.23 |
| ***APOE4*: 1 HDL-C: 0.6** | 2 | 13.86 | 10.50, 17.21 |
| | 4 | 14.48 | 12.14, 16.83 |
| | 8 | 15.73 | 12.97, 18.50 |
| | 12 | 16.99 | 11.70, 22.27 |
| ***APOE4*: 1 HDL-C: 2** | 2 | 15.09 | 11.78, 18.41 |
| | 4 | 16.05 | 13.92, 18.17 |
| | 8 | 17.96 | 15.20, 20.71 |
| | 12 | 19.86 | 14.20, 25.53 |

| | | | |
|---|---|---|---|
| ***APOE4*: 1 HDL-C: 3.3** | 2 | 16.24 | 8.61, 23.87 |
| | 4 | 17.50 | 13.39, 21.61 |
| | 8 | 20.02 | 14.02, 26.02 |
| | 12 | 22.54 | 8.72, 36.35 |
| ***APOE4*: 2 HDL-C: 0.6** | 2 | 14.82 | 8.29, 21.35 |
| | 4 | 15.56 | 11.54, 19.58 |
| | 8 | 17.03 | 11.56, 22.51 |
| | 12 | 18.51 | 6.99, 30.03 |
| ***APOE4*: 2 HDL-C: 2** | 2 | 15.71 | 9.49, 21.93 |
| | 4 | 17.11 | 13.68, 20.54 |
| | 8 | 19.90 | 14.76, 25.04 |
| | 12 | 22.70 | 11.19, 34.20 |
| ***APOE4*: 2 HDL-C: 3.3** | 2 | 16.54 | 1.01, 32.07 |
| | 4 | 18.55 | 10.75, 26.35 |
| | 8 | 22.57 | 9.88, 35.25 |
| | 12 | 26.59 | -3.00, 56.17 |

**Supplementary** Table 3. Subgroup analysis of glucose, HDL-C, and *APOE4* association with HCI for individuals with AD

| Variable | Coefficient | 95% CI | p-value |
|---|---|---|---|
| Glucose | 0.71 | -0.26, 1.67 | 0.15 |
| HDL-C | 0.92 | -2.12, 4.11 | 0.55 |
| *APOE4* | -3.05 | -5.92, -0.20 | 0.04* |
| Race (White) | 3.60 | -4.19, 11.40 | 0.37 |
| Visits | 2.90 | 2.12, 3.73 | 0.000*** |

Note. $^{*}p<0.05$; $^{***}p<0.001$

**Supplementary** Table 4. Subgroup analysis of glucose, HDL-C, and *APOE4* association with HCI for individuals without AD

| Variable | Coefficient | 95% CI | p-value |
|---|---|---|---|
| Glucose | 0.31 | 0.01, 0.60 | 0.04* |
| HDL-C | 1.33 | 0.27, 2.38 | 0.01* |
| *APOE4* | 1.72 | 0.75, 2.68 | 0.000*** |
| Race (White) | 3.17 | 0.73, 5.60 | 0.01* |
| Visits | 1.27 | 0.97, 1.58 | 0.000*** |

Note. *p<0.05; ***p<0.001
